# INCORPORATION OF NON-INVASIVE, CONTRAST-FREE, AND MINIMAL RADIATION PARAMETERS IN THE DIAGNOSTIC MODALITIES OF CORONARY ARTERY DISEASE

**DOI:** 10.1101/2024.12.04.24318528

**Authors:** Ibrahim Gul, Naqeeb Ullah Khan, Muhammad Wasim Awan, Naila Nasir Usmani, Mahjabeen Mahmood Kamal

## Abstract

**Background:** Ischemic heart disease is the major cause of morbidity and mortality. The diagnosis of chronic coronary syndrome is a challenge as most of the diagnostic investigation are either invasive or involve radiation or contrast material. The aim of our study was to find a scoring system comprising diagnostic parameters which are non-invasive and involve minimal radiation or contrast material and yet have high diagnostic yield.

**Methods:** In this non-randomized study conducted at KRL hospital Islamabad from Oct 2022 to Aug 2024, we took baseline characteristics and calculated Non Invasive, Minimal Radiation score (NIMR score) comprising risk factors, ETT, coronary calcium score and age of the patients, aged 18 and above. They then underwent CCTA to look for CAD. The association of the CAD with the score was checked using regression analysis on IBM SPSS version 26.

**Results:** Out of total 150 participants, 60% were male, mean age with SD was 50.54±7.82 and age ranged from 40-67 years. CAD was more prevalent in male and increased with increasing age. On Univariate analysis, NIMR score both as continuous variable and as categorical variable emerged as strong predictor of CAD (OR: 2.45, 95% CI: 1.84-3.25, pValue: 0.000) and (OR: 64.09, 95% CI: 14.53-282.74, pValue: 0.000) respectively. On multivariate analysis also the NIMR score was strong predictor of CAD both as continuous variable (OR: 2.43, 95%CI: 1.83-3.24, pValue: 0.000) and as categorical variable (OR: 67.11, 95%CI: 14.47-311.14, pValue: 0.000).

**Conclusion:** In conclusion, NIMR score, which is non-invasive, with minimal radiation and no contrast material involvement is a strong predictor of CAD in low and intermediate risk patients and very applicable in day to day clinical practice.

## INTRODUCTION

Ischemic heart disease (IHD) is the leading cause of morbidity and mortality worldwide.(1) Amongst the cardiovascular illnesses, IHD is considered the most prevalent of all, responsible for one third of all deaths worldwide and is acknowledged as the biggest threat to sustainable development of the 21^st^ century.(2, 3) The situation in lower and middle income countries like ours is grave and is expected to get worse due to the rise in aging population, urbanization and globalization with resultant shift in disease related morbidity and mortality from communicable diseases to non-communicable ones.(4) There is no single cause of IHD to be concentrated on and multiple modifiable and non-modifiable risk factors have been proposed which interplay in various amount to result in an ischemic event.(3)

IHD is classified as acute coronary syndrome (ACS) and chronic coronary syndrome (CCS). The assessment, diagnosis and management of ACS is straightforward in most of the cases and almost all cases ultimately need coronary angiography (CA) with view to revascularization either by angioplasty or coronary artery bypass grafting (CABG).(5) The difficulty in investigation arises in suspected CCS patients because simple test like Electrocardiography, Echocardiography, and Exercise tolerance tests (ETT) have very minimal diagnostic yield and cardiac biomarkers are negative. While many other more advanced diagnostic investigations for such stable/CCS patients have been developed as for example Single Photon Emission Computed Tomography (SPECT) Myocardial Perfusion Scan (MPS), Positron Emission Tomography (PET) scan, Coronary CT Angiography (CCTA) and Invasive Coronary Angiography (ICA) etc. they often suffer from either low sensitivity and specificity or limited patient acceptability due to the involvement of radiation and contrast materials or they are invasive.(6, 7) Stress echocardiography (SE), although is free of radiation and contrast material exposure, with sensitivity and specificity comparable to SPECT MPS but its interpretation is totally subjective, the images in case of exercise stress echocardiography need to be taken in a very limited time frame, there is problem of appropriate echocardiographic windows in some patients and is thus less preferable in many centers and to many patients.(8) Coronary Artery Calcium Scoring (CACS) has been extensively studied and is strongly associated with CAD. But solely depending on CACS would be in-appropriate.(9) Research suggests that combining ETT with CACS (referred to as Calcium Stress Testing) can enhance diagnostic accuracy.(10) The current gold standard for diagnosing coronary artery disease (CAD) is ICA. However, it is expensive and invasive, involves significant radiation exposure, the use of contrast material, and limited accessibility.(6, 11) Also ICA lacks the assessment of the functional significance of the stenotic lesion and some functional studies like Fractional Flow Reserve (FFR) or Instantaneous-wave Free Ratio (iFR) etc. need to be combined with it, in some situations, for appropriate conclusion to be drawn.(7)

The issue of choosing an appropriate diagnostic test in Non ACS setting, especially in intermediate risk CAD patients is paramount because, on one hand, the selected diagnostic modality should be having reasonable sensitivity and specificity, owing to the life threatening nature of the disease, while on the other hand, the patients should not unnecessarily be exposed to radiation, contrast medium or to invasive tests. The current societal guidelines also recommend some kind of non-invasive tests in such patients before embarking on invasive diagnostic procedures.(12) That is why need has always been felt to find out some novel diagnostic tests or devise some methods to diagnose CAD in such patients with no or less radiation, no contrast material and high accuracy.

Thus this study aims to develop a comprehensive scoring system that incorporates patient age and other risk factors, ETT data, and CACS to improve CAD diagnostic accuracy. The goal is to create a non-invasive, cost-effective diagnostic tool with minimal radiation exposure and no contrast material, making it accessible and acceptable for all segments of population.

## METHODS

This is a non-randomized, single-arm interventional study that was conducted at KRL Hospital, Islamabad from Oct 2022 to Aug 2024. The study was approved by the Ethical Review Committee of KRL hospital Islamabad (KRL-HI-PUB-ERC/Oct22/19) and was conducted according to the Good Clinical Practice Guidelines. KRL organization is a federal organization, having employees from all over the country including Gilgit Baltistan and Kashmir. Moreover, The major Medical Services Division (MSD) of KRL is in Islamabad and thus the study population form KRL hospital Islamabad is truly representative of the whole country. Written informed consent was taken from all the eligible candidates prior to enrollment into the study. The study population included patients aged 18 years and older who were being electively investigated for the possibility of CAD. As a part of the non-invasive assessment including history, physical examination, Electrocardiography, Echocardiography, laboratory testing, they also underwent ETT to assess myocardial ischemia. Patients who’s Duke Treadmill Score (DTS) indicated low or intermediate risk for CAD, underwent CT coronary artery calcium scoring via V-score Software, after providing informed consent as per our department protocol. Patients with a high-risk DTS were excluded from the study and they were straightaway referred for ICA. Also patients with acute coronary syndrome, a history of prior Percutaneous Coronary Intervention (PCI), prior CABG, chronic kidney disease (CKD), end-stage renal disease (ESRD), hyperparathyroidism, and malignancy were excluded from the study.

The ETT was conducted by a qualified cardiologist following the Bruce protocol, and the DTS was calculated using the following formula:

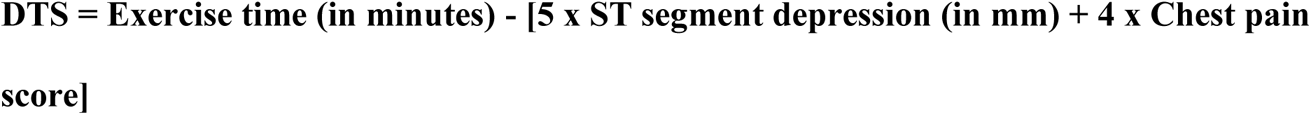

Chest pain score was as follows:

- 0: No chest pain
- 1: Chest pain not limiting exercise
- 2: Chest pain limiting exercise

DTS was classified as:

- Low risk: 0 points
- Intermediate risk: 2 points

CACS was performed by a trained radiologist, blinded to the patient’s clinical data. The scoring was categorized and scored as:

- No calcium: 0 points
- 1-100 calcium: 2 points
- 101-400 calcium: 4 points
- More than 400 calcium: 6 points

Age was classified and scored as follows:

- <40 years: 0 points
- 41-50 years: 1 points
- 51-60 years: 2 points
- 61-70 years: 3 points

Risk factors such as hypertension (HTN), diabetes mellitus (DM), dyslipidemia (DLP), family history, and smoking were each assigned 1 point.

### Final Scoring System

The total score was classified as follows:

- Low risk for CAD: 0-5
- Intermediate risk for CAD: 6-10
- High risk for CAD: 11-16

### Scoring Parameters

Scoring Parameters:
| Parameter | Score | Minimum Score | Maximum Score |
| --- | --- | --- | --- |
| **DTS** | Low risk: 0<br>Intermediate risk: 2 | 0 | 2 |
| **Coronary Calcium Score** | 0: 0<br>1-100: 2<br>101-400: 4<br>>400: 6 | 0 | 6 |
| **Risk Factors** | No risk factors: 0<br>HTN: 1<br>DM: 1<br>DLP: 1<br>Family history: 1<br>Smoking: 1 | 0 | 5 |
| **Age** | ≤40 yrs.: 0<br>41-50 yrs.: 1<br>51-60 yrs.: 2<br>61-70 yrs.: 3 | 0 | 3 |

The study subjects then underwent CCTA after taking written informed consent as per the protocol of our department and CCTA was assessed by a trained radiologist, who was also blind to the patient’s clinical data as discussed previously. CCTA was done with 640 slice CANON AQUILION machine, with slice thickness of 0.5mm. All the patients with heart rate more than 70 bmp were given beta blockers and or calcium channel blocker to control their heart rate. Each patient was given sublingual nitroglycerine 0.5 mg 1-2 minutes before taking the images. Non-ionic, low osmolar contrasts material was used as 1.5 ml/kg body weight in a bolus form at a rate of 6 ml/sec followed by saline flush of 50 ml. ECG recording was done for data reconstruction. Images were taken at 75% of the R-R interval. The mean radiation dose in single study was 400 mGy.cm. The gold standard in our study was CCTA and not ICA due to the following reasons.

1. The CCTA with 640 slice CT scanner with slice thickness of 0.5mm has accuracy more than 98.91% of the ICA.
2. The majority of the patients in our study were young and thus non-invasive procedure was more acceptable to them than the invasive procedure.
3. Due to being comparatively young study population, we also expected coronary anomalies and myocardial bridging which are better detected by CCTA than ICA.
4. CCTA has been used instead of ICA in some other major trials.(13)

The data was collected on a paper based questionnaire. All the authors contributed to study design, data collection, analysis, interpretation and writing the manuscript in one form or the other and all authors vouch for the accuracy and completeness of the data and analysis.

### Statistics

Data analysis was done using IBM SPSS version 26. Descriptive statistics were calculated for all the variables in the study. Means with standard deviations were calculated for continuous variables like age whereas proportions were calculated for categorical variables like HTN, DM etc. Cross tabulation was used to assess relationship between various variables like age and CAD and gender and CAD. Logistic regression analysis was used to take the final inference of association of NIMR score with CAD. All the 6 assumptions were first tested and found satisfying before binary logistic regression analysis was done. After that Univariate logistic regression analysis and then Multivariate logistic regression analysis were done to deduce the final conclusion of association of NIMR score with CAD. The details of each statistical step is written in the results section. A p-value <0.05 was considered statistically significant at the multivariate level. The final model was checked for outlying or influential observations and was reported with the overall log likelihood.

## RESULTS

During our study period, we investigated 180 subjects. Of these 180 subjects, 20 were excluded because their DTS was in high risk category and they were referred for ICA. Of the remaining 160 subjects, 5 were excluded due to CKD (2), Prior PCI (2) and Prior CABG (1). Five (5) more patients lost to follow for CCTA. Thus the final analysis was done on 150 individuals, consisting of 90 males (60%) and 60 females (40%).

Baseline characteristics of the study participants are given in Table 1.

**Table 1:** Baseline characteristics of the study participants.

| **Variables** | **No. Male** | **No. Female** | **Means±SD** | **No. (%)** |
| --- | --- | --- | --- | --- |
| *No of participants* | 90 | 60 |  |  |
| *Age yrs.* |  |  | 50.54±7.82 |  |
| 35-44 | 31 | 13 |  | 44(29.3) |
| 44-54 | 32 | 24 |  | 56(37.3) |
| 55-64 | 18 | 22 |  | 40(26.7) |
| 65-74 | 9 | 1 |  | 10(6.7) |
| *Hypertensive* | 52 | 29 |  | 81(54) |
| *Diabetics* | 18 | 7 |  | 25(16.7) |
| *Dyslipidemics* | 21 | 11 |  | 32(21.3) |
| *Family history positive* | 15 | 12 |  | 27(18) |
| *Smokers* | 9 | 2 |  | 11(7.3) |
| *Exercise tolerance Test* |  |  |  |  |
| Low risk | 28 | 28 |  | 56(37.3) |
| Intermediate risk | 62 | 32 |  | 94(62.7) |
| *Coronary Calcium score* |  |  |  |  |
| 0 | 38 | 51 |  | 89(59.3) |
| 1-100 | 30 | 7 |  | 37(24.7) |
| 101-400 | 14 | 0 |  | 14(9.3) |
| >400 | 8 | 2 |  | 10(6.7) |
| *Pattern of CAD* |  |  |  |  |
| Normal | 30 | 34 |  | 64(42.7) |
| SVCAD | 20 | 11 |  | 31(20.7) |
| DVCAD | 18 | 4 |  | 22(14.7) |
| TVCAD | 15 | 0 |  | 15(10.0) |
| Coronary anomaly | 0 | 9 |  | 9(6.0) |
| Myocardial Bridging | 7 | 2 |  | 9(6.0) |
| *Name of the Artery involved* |  |  |  |  |
| LAD | 24 | 12 |  | 36(24.0) |
| LCX | 0 | 2 |  | 2(1.3) |
| RCA | 3 | 8 |  | 11(7.3) |
| LAD/RCA | 4 | 1 |  | 5(3.3) |
| LAD/LCX | 14 | 3 |  | 17(11.3) |
| LAD/LCX/RCA | 15 | 0 |  | 15(10.0) |
| *Creatinine* |  |  | 0.88±0.17 |  |
| *Urea* |  |  | 29.26±6.58 |  |
| *Sodium* |  |  | 137.87±1.84 |  |
| *Potassium* |  |  | 3.93±0.23 |  |
| *Platelets* |  |  | 275.64±61.63 |  |

The mean age for both sexes combined was 50.54 years, with a standard deviation of 7.82 years. The participants’ ages ranged from a minimum of 40 years to a maximum of 67 years. (Supplementary table 1 and Supplementary Figure 1) We observed a clear trend in our data with respect to the participant’s age: as the age group increased, the number of individuals with CAD also increased in comparison to those without the disease within the same age group. This suggests that older age groups have a higher prevalence of CAD. (Supplementary table 2) Table 2 presents a cross-tabulation of the patient’s sex and the pattern of CAD. The data highlights that most forms of CAD were more prevalent in males compared to females, with the exception of coronary anomalies. All 9 cases of coronary anomalies were found exclusively in females. In terms of single vessel coronary artery disease (SVCAD), the incidence in males was nearly double that in females (20 cases in males vs. 11 in females). The disparity was even more pronounced for double vessel CAD (DVCAD), where males accounted for over four times the cases compared to females (18 vs. 4). Additionally, triple vessel CAD (TVCAD) was solely observed in males, with all 15 reported cases occurring in this group. The male-to-female ratio for myocardial bridging was 7:2.

**Table 2:** Cross tabulation of Sex of the Patient and Pattern of CAD.

| Pattern of CAD | Sex of the Patient |  | Total |
| --- | --- | --- | --- |
|  | Female | Male |  |
| Normal | 34 | 30 | 64 |
| SVCAD | 11 | 20 | 31 |
| DVCAD | 4 | 18 | 22 |
| TVCAD | 0 | 15 | 15 |
| Coronary Anomaly | 9 | 0 | 9 |
| Myocardial Bridging | 2 | 7 | 9 |
| Total | 60 | 90 | 150 |

Binary logistic regression was applied after confirming all the following necessary six assumptions to be true:

1. **The dependent variable must be a binary variable.** In our data CAD_Status is a binary variable with 0 = Normal and 1 = Diseased.
2. **Observations should be independent of each other.** In our data each patient was an independent observation where there were no repeated measures or grouped data.
3. **There must be no perfect multicollinearity among independent variables.** The Variance Inflation Factor (VIF) for **NIMR score & Sex** was 2.26, and for **NIMR score_Group**, it was 2.75, both values being below 5, suggesting no serious multicollinearity issues. Additionally, the Tolerance scores were 0.44 for **NIMR score & Sex** and 0.36 for **NIMR score_Group**, both comfortably exceeding the 0.2 threshold, confirming that multicollinearity is not a problem in our analysis.
4. **Linearity:** As we have only one continuous variable (NIMR score), we plotted the log-odds (logit) of the dependent variable, **CAD_Status**, against **NIMR score**. The plot reveals a perfect linear relationship between the two variables as shown in Figure 1.
5. **Sufficient sample size.** Logistic regression requires a sufficiently large sample size to generate reliable estimates. A widely accepted guideline is the 10 events per variable (**EPV**) rule **6**,(14) which suggests that for each independent variable, there should be at least 10 events (in this case, cases of CAD). Our dataset consists of 150 observations, including 86 individuals with CAD (Diseased = 1) and 64 without CAD (Normal = 0). Given that we are using two independent variables, the data comfortably satisfies the 10 EPV rule, ensuring that the model is well-suited for robust and accurate analysis.
6. **Absence of Outliers or Influential Data Points.** To identify potential outliers or influential observations, we conducted Cook’s Distance test. In our analysis, all Cook’s Distance values were below 1, confirming that none of the individual data points are significantly influencing the outcome of the logistic regression model.

**Figure 1:**
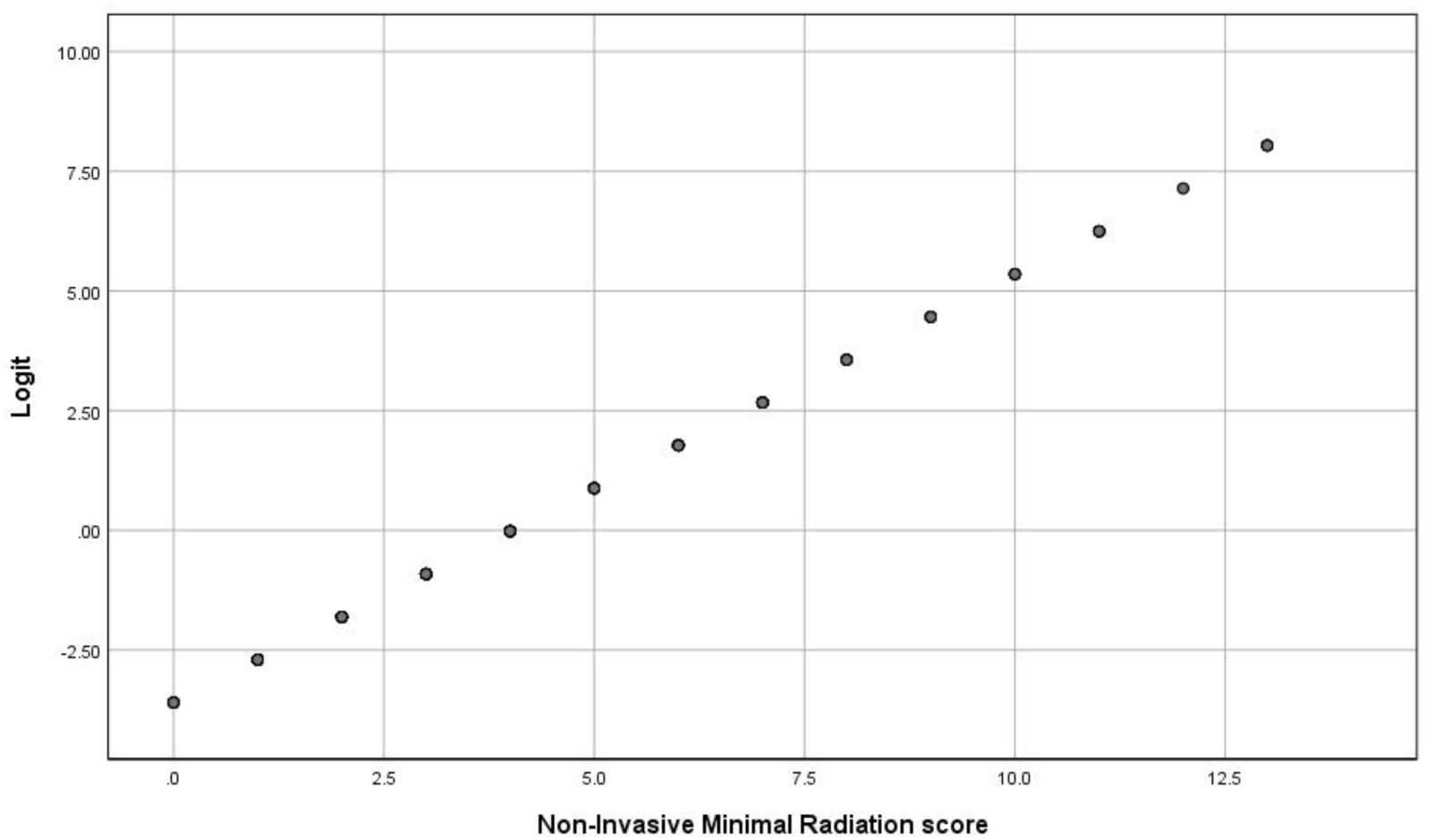
Linear relationship between NIMR score & CAD_Status.

### Univariate Logistic Regression Analysis

The results of the Univariate logistic regression analysis for the independent variables in our study—**Sex**, **NIMR score**, and **NIMR score_Group**—are presented in Table 3. Each of these variables showed a significant association with the dependent variable, **CAD_Status**. Males were found to be at a considerably higher risk of developing CAD compared to females, with an odds ratio (OR) of 2.62 and a 95% confidence interval (CI) ranging from 1.33 to 5.13

**Table 3:** Table: Univariate analysis of the predictors with the CAD_Status.

| Variable | Coefficients | OR | p-value | 95% CI |  |
| --- | --- | --- | --- | --- | --- |
|  |  |  |  | Lower | Upper |
| Sex | 0.96 | 2.62 | 0.005 | 1.33 | 5.13 |
| NIMR score | 0.90 | 2.45 | 0.000 | 1.84 | 3.25 |
| NIMR Score_Group | 4.16 | 64.09 | 0.000 | 14.53 | 282.74 |

**Figure 2** shows a graph, in which there is a clear upward trend: as the **NIMR score** increases, the log-odds of having CAD also increase. This means that individuals with higher NIMR scores are significantly more likely to have CAD. The linear pattern in the log-odds suggests a strong positive relationship between the NIMR score and the likelihood of developing CAD.

**Figure 2:**
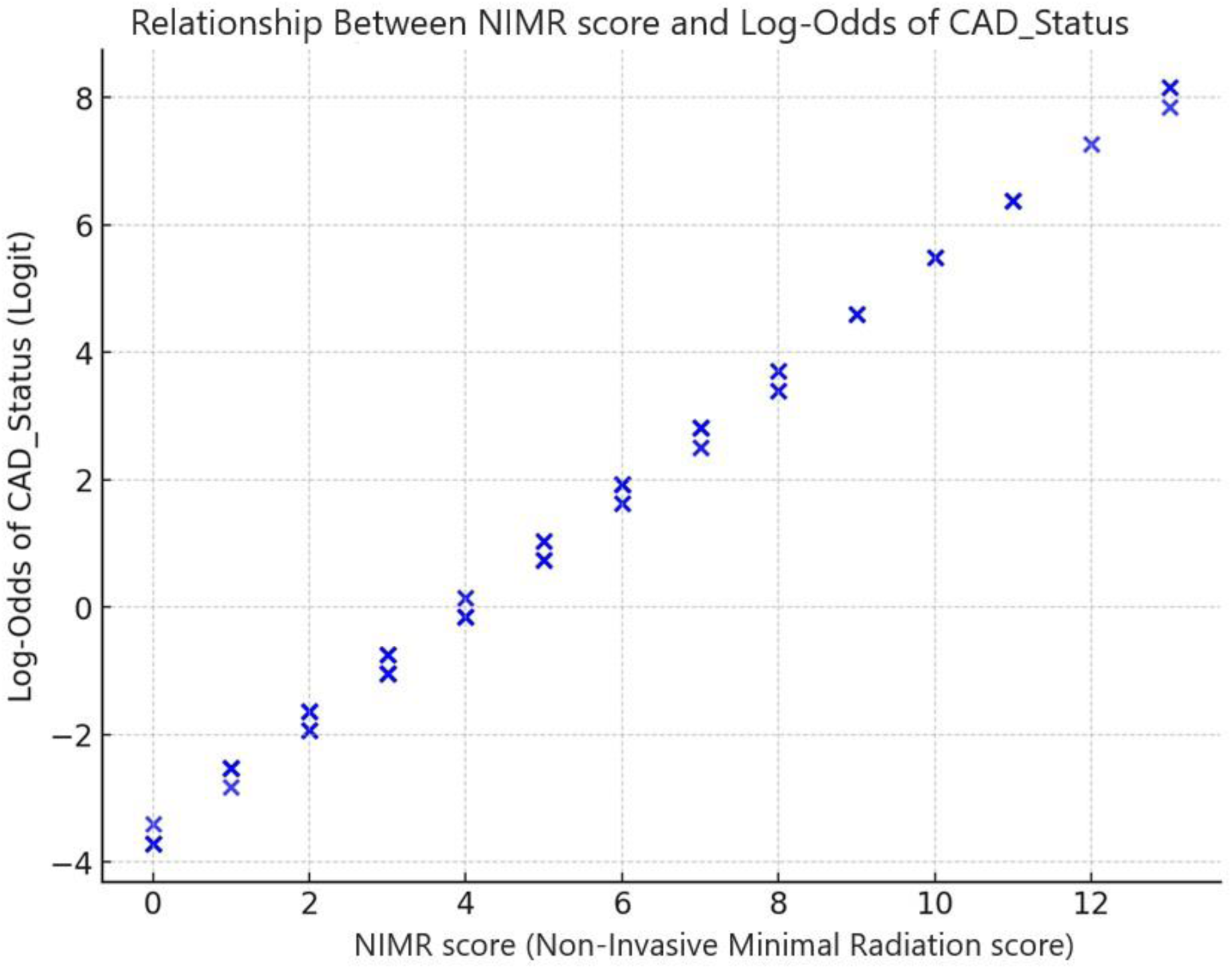
Relationship between NIMR score and CAD_Status.

### Multivariate Logistic Regression Analysis

The results of the multivariate logistic regression, along with the adjusted odds ratios for the variables associated with CAD, are presented in Table 4. In this model, the predictors are **Sex** and the **NIMR score**, with **CAD_Status** as the outcome variable.

**Table 4:** Multivariate logistic regression analysis of factors associated with CAD.

| Variables | Coefficient | p-Value | OR | 95% C.I for OR |  |
| --- | --- | --- | --- | --- | --- |
|  |  |  |  | Lower | Upper |
| NIMR score | 0.89 | 0.000 | 2.43 | 1.83 | 3.24 |
| † Sex | 0.31 | 0.542 | 1.36 | 0.51 | 3.64 |
| †- Female Sex was taken as a reference category |  |  |  |  |  |

In our analysis, **Sex** was not a significant predictor, as there was no strong evidence suggesting that males were more likely to have CAD than females after controlling for NIMR score. On the other hand, **NIMR score** emerged as a significant predictor, with an odds ratio (OR) of 2.43. Both the p-value and 95% confidence interval (CI) for NIMR score were highly significant, indicating that for every unit increase in NIMR score, the odds of having CAD increase by approximately 2.43 times when adjusting for other variables. In simple words we can say that a higher NIMR score is a strong risk factor for CAD.

The figure below clearly illustrates the robust relationship, demonstrating how an increase in the **NIMR score** correlates with a higher likelihood of developing CAD. This visual representation reinforces the statistical findings, showing a clear upward trend in the risk of CAD as the NIMR score increases.

**Figure 3:**
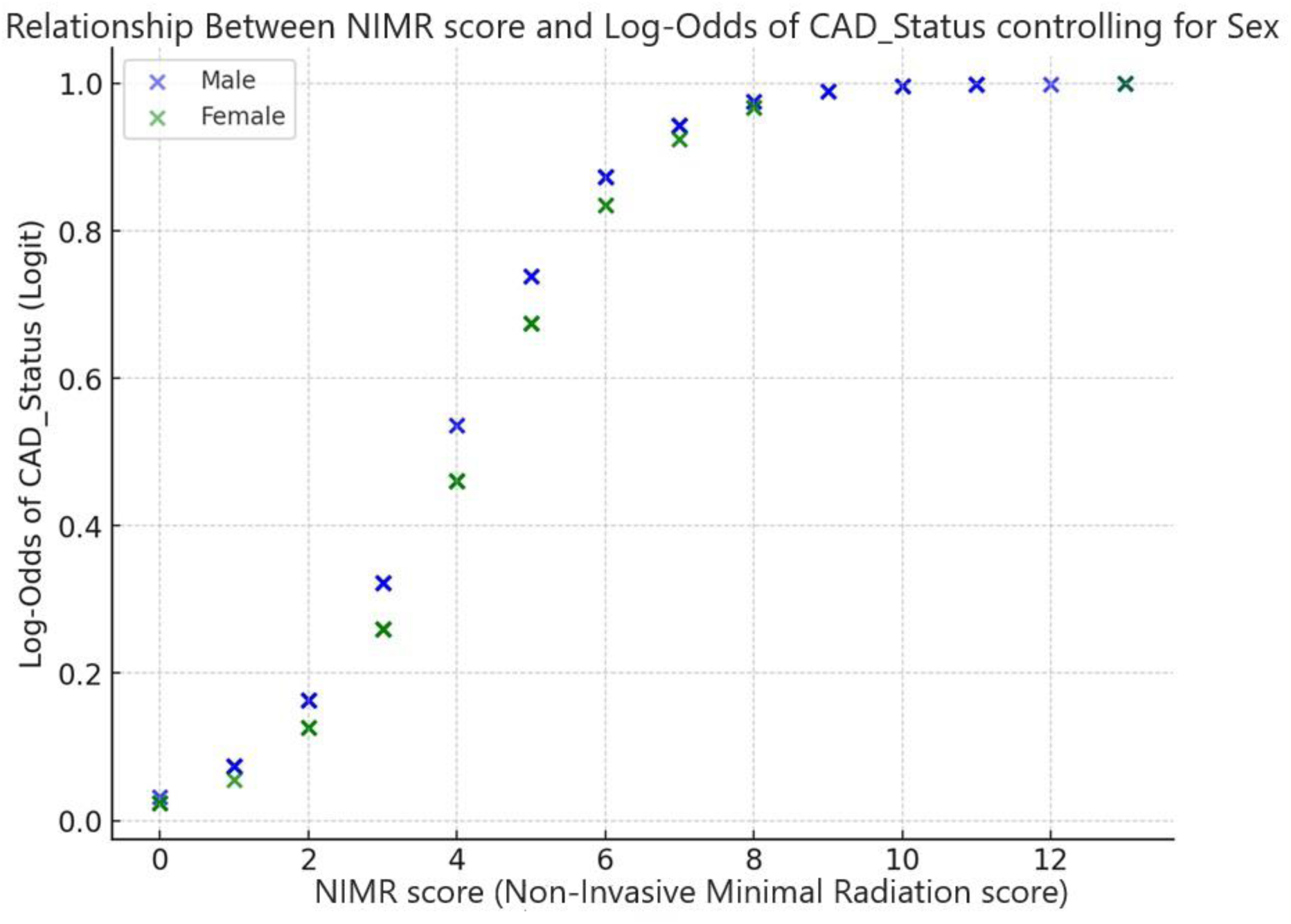
Relationship between NIMR and CAD _Status.

In this study, we began by performing a logistic regression using **NIMR score** as a continuous variable alongside **Sex** to predict **CAD_Status**. To enhance the analysis, we categorized the **NIMR score** into three distinct risk groups, creating a new variable called **NIMR score_Group**. This approach mirrors clinical practice, where risk factors are typically grouped into meaningful thresholds, such as low, intermediate, and high risk, to reflect varying levels of disease risk. Also by treating **NIMR score** as a categorical variable, we aimed to capture potential non-linear relationships between the risk score and **CAD_Status** that may not be evident when using it as a continuous variable.

Table 5 presents the logistic regression results for the model where **Sex** and **NIMR score_Group** (with the first category as the reference group) are the independent variables, and **CAD_Status** is the dependent variable. The results are summarized as follows:

- **NIMR score_Group** (**2**): The odds ratio (OR) is 67.11, with a significant p-value and 95% confidence interval (CI), indicating that patients in the second risk group have significantly higher odds of having CAD compared to those in the reference group (NIMR score_Group 1).
- **NIMR score_Group** (**3**): The odds ratio in this group is extremely large, likely due to the high coefficient value, possibly a result of quasi-separation. In this study, all 16 subjects in the high-risk group (NIMR score_Group 3) had CAD, with none being healthy.
- **Sex**: With females as the reference category, the p-value and 95% CI for the odds ratio were not statistically significant. This suggests that, after accounting for risk categories, there is no significant difference in the odds of having CAD between males and females.

**Table 5:** Multivariate logit analysis of factors associated with CAD when NIMR score_Group *was taken as a categorical variable*.

| Variables |  | Coefficients | pValue | OR | 95% C.I.for OR |  |
| --- | --- | --- | --- | --- | --- | --- |
|  |  |  |  |  | Lower | Upper |
| †NIMR score_Group | NIMR score_Group (2) | 4.21 | 0.000 | 67.11 | 14.47 | 311.14 |
|  | NIMR score_Group (3) | 22.27 | 0.998 | 4684107184.40 | 0.00 |  |
| ‡Sex |  | -0.16 | 0.730 | 0.84 | 0.33 | 2.16 |
| † NIMR score_Group (1) was selected as a reference group. ‡ Female Sex was selected as a reference category |  |  |  |  |  |  |

For complete statistical details of the results, see (Supplementary file 2).

## DISCUSSION

As discussed initially, CAD is dual challenge to the world, both in terms of morbidity and mortality amongst the non-communicable diseases and in terms of the diagnostic dilemma of CAD because almost all the diagnostic modalities which bear the potential of definitive diagnosis are either invasive or expensive or carry the hazard of radiation and contrast material exposure.(7, 12). That is why intense need has always been felt to device some appropriate scoring systems or to find out some diagnostic tests to fill the gap in the diagnostic armamentarium of CAD. Our study in this respect is very important because we have devised a scoring system (NIMR score) for patients with intermediate risk CAD based on ETT, which help reclassify such patients in to low risk, intermediate risk and high risk categories so that many patients can be safely excluded from exposure to high dose radiation or contrast medium and only those patients who most likely need some intervention are subjected to invasive procedures. Our score has linear relation with the CAD such that the higher the score, the more are chances that the patient is having underlying significant CAD. Our NIMR score has significant linear relation both as continuous variable and as categorical i.e. when the score is classified into low, intermediate and high risk groups. This is important because such classification is easy to apply in day to day clinical practice.

Various algorithms and scores have been developed previously in such patients which have different sensitivity and specificity, applicable in different patients groups and in different geographical locations, as for example Framingham Risk Score (FRS) to predict 10 years risk of cardiovascular events, PROCAM for acute myocardial infarction, SCORE algorithm to assess death due to myocardial infarction and Diamond and Forrester pre-test probability tables etc.(15) The Framingham study, one of the landmark study and the FRS for 10 years risk of cardiovascular events is of immense importance in the field of preventive cardiology.(16) FRS which is the initial screening tool, is based on history, clinical examination and laboratory measurements without any stress testing and some important risk factors, which is primarily meant for asymptomatic patients. That is why it fails to identify many peoples who later on develop serious cardiovascular events.(17) Our score is different because it is for the clinical practice of day to day patients who present to OPD with suspected CAD and who need assessment to decide whether to go for invasive work up or not. Thus patients with high NIMR score can be referred to ICA, those with low risk can be safely discharged home and those with intermediate risk can be further tested with Either CCTA or SPECT MPS or SE. Our score has almost all the common and important risk factors including age, the stress testing and the CACS and thus it is a comprehensive and yet very less expensive and safe which very smartly re-categorize the most ambiguous group of patients i.e. ETT based low and intermediate risk groups and help in appropriately deciding their fate in terms of further management.

CACS has been widely studied and has been shown to be significantly associated with CAD especially in asymptomatic patients but also in symptomatic patients.(18) Also the Framingham study which is ongoing project has suggested strong association of CACS with CAD. But CACS in isolation might not be that strongly associated with CAD than when it is assessed in conjunction with other risk factors. Our NIMR score has the beauty that it has combined CACS along with other almost all important risk factors as well as stress testing and that is probably the reason we have found robust association of our score with CAD.

Diamond and Forrester probability score has been widely used, updated and tested to be appropriately applied for low and intermediate risk patients in clinical practice. Its revalidation suggested that adding stress testing would not add to the diagnostic accuracy of the score. However they just tested symptoms, age and gender of the patients.(19) Also this pre-test probability algorithm requires some additional testing like CCTA etc. to overcome its overestimation of CAD in low prevalent regions like Mediterranean countries, as recently reported.(20) Our NIMR score is different in that we have incorporated all the common risk factors, calcium score, ETT and age of the patients and thus it more accurately and more intensely investigate such category of patients. That is the reason our NIMR score has strong correlation with CAD. Also it classify patients into risk groups so that an appropriate and immediate decision can be taken for further management of such patients without much need of incorporation of battery of additional investigation.

European Society of Cardiology pre-test probability algorithm has incorporated both risk factors, CACS and other test and classified the CCS patients into high likelihood and low likelihood groups which too is a comprehensive algorithm that has good applicability to day to day clinical practice.(21) However NIRM score is different in that ECG and Echocardiography have not been incorporated into it. It is because, in patients with low and intermediate risk groups on the basis of ETT, these two tests are almost always normal and that is why such patients undergo ETT. If patient who present to OPD with chest pain and ECG or echocardiographic abnormalities, such patients usually do not need stress testing and they are directly referred to ICA. Our score is especially meant to reclassify the very stable patients with unremarkable initial investigation including ECG and Echocardiography and still has the potential to be at risk for CAD.

Our NIMR score has unique importance in that it has got very minimal investigation as well as minimal radiation and no contrast material. The applicability in resource restricted settings like ours which also is at the high end of the graph of CAD prevalence, is most appropriate. Our society on one hand is resource-restrained and on the other hand, we have very high burden of CAD due to changing population demographics, westernization of our society and the genetic predisposition.(22) We therefore cannot afford the expensive investigation as well as investigation carrying radiation hazards with late expected rise in carcinogenesis. That is why NIMR score is best suited for resource limited regions, to exclude part of the patient bulk from subjecting to expensive tests. Also due to very minimal radiation of the CACS and no contrast material involvement, affluent societies can also adopt this score to avoid the hazards of radiation and contrast material. Also this score will be useful for those patients with deranged renal function and for those subjects who are allergic to contrast material.

Apart from the NIMR score, there are some other interesting findings in our study which need to be discussed. In our study, the prevalence of coronary artery disease increases as the age of the study participants increases. This is in accordance with the established research and the understood fact that age i.e. “simply getting older” is a non-modifiable risk factor for CAD.(23)

In our study, the prevalence of CAD is significantly higher in male participants than female participants. Not only this but the association is stronger as the number of the involved coronary arteries increases such that all patients with TVCAD are male and the number of female with DVCAD are also far less than male participants. This is in accordance with the fact that CAD is more prevalent in male than female and male gender is an established risk factor for CAD. (12, 24)

We came across 6% of coronary anomalies in our study. Although variable percentages have been quoted in different studies, the one most authentic paper is by Angelini and his coworkers,(25) who thoroughly evaluated 1950 consecutive cineangiograms and quoted 5.6% prevalence of coronary anomalies. It means that our data is concordant with the true prevalence of coronary anomalies. One reason why we found out the exact percentage might be that we used CCTA instead of ICA as coronary anomalies may be missed during ICA which is just luminogram with little special orientation as compared to CCTA. In our study, Right Coronary Artery (RCA) anomaly was the most common one. Studies differ with respect to the type of the artery having anomaly. Some studies quote LCX artery anomaly to be the most common one whereas some others quote differently.(26) This might be because, the total number of anomalous coronary arteries in our study is very less. Large data may verify the true percentages according to the international prevalence. Interestingly, all the coronary anomalies were found in female participants. International research also support our finding as is evident from the studies that coronary anomalies are more common in female population.(27)

In our study, we found 9 cases of myocardial bridging with 7:2 male to female preponderance. This accounts for 6% of our study participants and most of the myocardial bridging was in Left Anterior Descending (LAD) artery. This is in accordance with the international data which quote very wide prevalence ranging from 0.15% to 25% and LAD is the most commonly involved artery in myocardial bridging.(28) One reason of the wide difference in the prevalence of both coronary anomalies and myocardial bridging might be that some studies quote both the coronary anomaly and myocardial bridging in a single heading of coronary anomalies whereas others report coronary anomalies different from myocardial bridging. We analyzed both of them differently. If we combine both in a single heading of coronary anomalies, then too, the prevalence percentage is as per the international data.

### Limitations of our study

The no of participants in high risk group of NIMR score are limited and all are having CAD which to some extent affects the statistical analysis by projecting very high Odd Ratio. Further studies with large no of sample are required to elucidate it. The gold standard in our study was CCTA. Further studies can be done with gold standard as ICA with FFR or iFR so that the results are revalidated along with functional assessment of the coronary stenosis. We did not incorporate gender in the risk score whereas the Diamond and Forrester probability score has gender as one of major components. Although we run the statistics to look as to whether gender has significant association and the results showed marginal significance after applying the NIMR score groups but still gender can be incorporated in to the score and reapply in such population.

### Strengths of our study

The CCTA was done with 640 slice CT scanner with slice thickness of 0.5 mm which has very high accuracy. The NIRM score was having linear relationship both as continuous as well as categorical variable and thus good applicability in clinical practice. The study population is the true representative of the whole country, being a single major MSD of the federal organization. The parameter in our score are all less expensive, with minimal radiation and no contrast material involved.

### Conclusion

In conclusion, NIMR score is a very practicable, less expensive, minimal radiation and contrast free score for low and intermediate risk CAD patients which can be applied in day to day clinical practice for both resource limited segments of population due to being cost effective and to affluent population due to being contrast free and with minimal radiation.

## Data Availability

All the data will be made available if and when required

## ACKNOWLEDGEMENT

We acknowledge the technical support, in setting, arrangement of medicine and performance of CCTA, of Mr Sharfuddin, Principal Technician-II, FSC 2 years diploma in radiology, Cell No:+92 321 5344224

## SOURCE OF FUNDING

No funding involved

## DISCLOSURE

None

## Notes

### Competing Interest Statement

The authors have declared no competing interest.

### Author Declarations

Ref ERC: KRL-HI-PUB-ERC/Oct22/19 Ethical Review committee of KRL Hospital Islamabad has thorougly reviewed the study of Dr Ibrahim Gul of this hospital Titled :INCORPORATION OF NON-INVASIVE, CONTRAST-FREE, AND MINIMAL RADIATION PARAMETERS IN THE DIAGNOSTIC MODALITIES OF CORONARY ARTERY DISEASE" which he will conduct in KRL hospital Islamabad. The committee did not find anything in this study which is unethical, injurious or against the international guidelines for biomedical research involving human or animal subjects.

## REFERENCES

1. Benjamin EJ, Blaha MJ, Chiuve SE, Cushman M, Das SR, Deo R, et al. Heart Disease and Stroke Statistics-2017 Update: A Report From the American Heart Association. Circulation. 2017;135(10):e146–e603.

2. Roth GA, Johnson C, Abajobir A, Abd-Allah F, Abera SF, Abyu G, et al. Global, Regional, and National Burden of Cardiovascular Diseases for 10 Causes, 1990 to 2015. J Am Coll Cardiol. 2017;70(1):1–25.

3. The changing patterns of cardiovascular diseases and their risk factors in the states of India: the Global Burden of Disease Study 1990-2016. Lancet Glob Health. 2018;6(12):e1339–e51.

4. Khan MA, Hashim MJ, Mustafa H, Baniyas MY, Al Suwaidi S, AlKatheeri R, et al. Global Epidemiology of Ischemic Heart Disease: Results from the Global Burden of Disease Study. Cureus. 2020;12(7):e9349.

5. Gulati M, Levy PD, Mukherjee D, Amsterdam E, Bhatt DL, Birtcher KK, et al. 2021 AHA/ACC/ASE/CHEST/SAEM/SCCT/SCMR Guideline for the Evaluation and Diagnosis of Chest Pain: Executive Summary: A Report of the American College of Cardiology/American Heart Association Joint Committee on Clinical Practice Guidelines. Journal of the American College of Cardiology. 2021;78(22):2218–61.

6. Albus C, Barkhausen J, Fleck E, Haasenritter J, Lindner O, Silber S. The Diagnosis of Chronic Coronary Heart Disease. Dtsch Arztebl Int. 2017;114(42):712–9.

7. Danad I, Raijmakers PG, Driessen RS, Leipsic J, Raju R, Naoum C, et al. Comparison of Coronary CT Angiography, SPECT, PET, and Hybrid Imaging for Diagnosis of Ischemic Heart Disease Determined by Fractional Flow Reserve. JAMA Cardiology. 2017;2(10):1100–7.

8. Pellikka PA, Arruda-Olson A, Chaudhry FA, Chen MH, Marshall JE, Porter TR, Sawada SG. Guidelines for Performance, Interpretation, and Application of Stress Echocardiography in Ischemic Heart Disease: From the American Society of Echocardiography. Journal of the American Society of Echocardiography. 2020;33(1):1–41.e8.

9. Liaquat A, Khan A, Ullah Shah S, Iqbal H, Iqbal S, Rana AI, Ur Rahman H. Evaluating the use of coronary artery calcium scoring as a tool for coronary artery disease (CAD) risk stratification and its association with coronary stenosis and CAD risk factors: a single-centre, retrospective, cross-sectional study at a tertiary centre in Pakistan. BMJ Open. 2022;12(7):e057703.

10. Ananthasubramaniam G, Ananthasubramaniam K. Stress electrocardiography testing in coronary artery disease: Is it time for its swan song or to redefine its role in the modern era? Indian Heart J. 2022;74(2):81–5.

11. Wardziak Ł, Kruk M, Pleban W, Demkow M, Rużyłło W, Dzielińska Z, Kępka C. Coronary CTA enhanced with CTA based FFR analysis provides higher diagnostic value than invasive coronary angiography in patients with intermediate coronary stenosis. J Cardiovasc Comput Tomogr. 2019;13(1):62–7.

12. Virani SS, Newby LK, Arnold SV, Bittner V, Brewer LC, Demeter SH, et al. 2023 AHA/ACC/ACCP/ASPC/NLA/PCNA Guideline for the Management of Patients With Chronic Coronary Disease. Journal of the American College of Cardiology. 2023;82(9):833–955.

13. Maron DJ, Hochman JS, Reynolds HR, Bangalore S, O’Brien SM, Boden WE, et al. Initial Invasive or Conservative Strategy for Stable Coronary Disease. New England Journal of Medicine. 2020;382(15):1395–407.

14. Hodge AM, English DR, O’Dea K, Giles GG. Dietary Patterns and Diabetes Incidence in the Melbourne Collaborative Cohort Study. American Journal of Epidemiology. 2007;165(6):603–10.

15. Versteylen MO, Joosen IA, Shaw LJ, Narula J, Hofstra L. Comparison of Framingham, PROCAM, SCORE, and Diamond Forrester to predict coronary atherosclerosis and cardiovascular events. Journal of Nuclear Cardiology. 2011;18(5):904–11.

16. Hemann BA, Bimson WF, Taylor AJ. The Framingham Risk Score: An Appraisal of Its Benefits and Limitations. American Heart Hospital Journal. 2007;5(2):91–6.

17. Brindle P, Beswick A, Fahey T, Ebrahim S. Accuracy and impact of risk assessment in the primary prevention of cardiovascular disease: a systematic review. Heart. 2006;92(12):1752–9.

18. Shreya D, Zamora DI, Patel GS, Grossmann I, Rodriguez K, Soni M, et al. Coronary Artery Calcium Score - A Reliable Indicator of Coronary Artery Disease? Cureus. 2021;13(12):e20149.

19. Sørgaard M, Linde JJ, Kofoed KF, Kühl JT, Kelbæk H, Nielsen WB, Hove JD. Diagnostic Value of the Updated Diamond and Forrester Score to Predict Coronary Artery Disease in Patients with Acute-Onset Chest Pain. Cardiology. 2016;133(1):10–7.

20. Marcos-Alberca P, De Agustin JA, Gomez De Diego JJ, Pozo-Osinalde E, Bustos A, Cabeza B, et al. Overestimation of the pretest probability of coronary heart disease with the Diamond and Forrester algorithm in low prevalent Mediterranean area. European Heart Journal - Cardiovascular Imaging. 2023;24(Supplement_1).

21. Knuuti J, Wijns W, Saraste A, Capodanno D, Barbato E, Funck-Brentano C, et al. 2019 ESC Guidelines for the diagnosis and management of chronic coronary syndromes: The Task Force for the diagnosis and management of chronic coronary syndromes of the European Society of Cardiology (ESC). European Heart Journal. 2019;41(3):407–77.

22. Sucato V, Coppola G, Manno G, Vadalà G, Novo G, Corrado E, Galassi AR. Coronary Artery Disease in South Asian Patients: Cardiovascular Risk Factors, Pathogenesis and Treatments. Current Problems in Cardiology. 2023;48(8):101228.

23. Hajar R. Risk Factors for Coronary Artery Disease: Historical Perspectives. Heart Views. 2017;18(3):109–14.

24. Dueñas M, Ramirez C, Arana R, Failde I. Gender differences and determinants of health related quality of life in coronary patients: a follow-up study. BMC cardiovascular disorders. 2011;11:1–11.

25. Angelini P, Villason S, Chan A, Diez J. Coronary artery anomalies: Baltimore: Lippincott Williams & Wilkins; 1999.

26. Kashyap JR, Kumar S, Reddy S, Rao KR, Sehrawat O, Kashyap R, et al. Prevalence and Pattern of Congenital Coronary Artery Anomalies in Patients Undergoing Coronary Angiography at a Tertiary Care Hospital of Northern India. Cureus. 2021;13(4):e14399.

27. Aydar Y, Yazici HU, Birdane A, Nasifov M, Nadir A, Ulus T, et al. Gender differences in the types and frequency of coronary artery anomalies. Tohoku J Exp Med. 2011;225(4):239–47.

28. Villa AD, Sammut E, Nair A, Rajani R, Bonamini R, Chiribiri A. Coronary artery anomalies overview: The normal and the abnormal. World J Radiol. 2016;8(6):537–55.

